# SARS-CoV-2 outbreaks in secondary school settings in the Netherlands during fall 2020; silent circulation

**DOI:** 10.1101/2022.05.02.22273861

**Authors:** Lotte Jonker, Kimberly J Linde, Marieke LA de Hoog, Robin Sprado, Robin C Huisman, Richard Molenkamp, Bas B Oude Munnink, Wietske Dohmen, Dick JJ Heederik, Dirk Eggink, Matthijs RA Welkers, Harry Vennema, Pieter LA Fraaij, Marion PG Koopmans, Inge M Wouters, Patricia CJL Bruijning-Verhagen

## Abstract

**Background:** In fall 2020 when schools in the Netherlands operated under a limited set of COVID-19 measures, we conducted outbreaks studies in four secondary schools to gain insight in the level of school transmission and the role of SARS-CoV-2 transmission via air and surfaces.

**Methods:** Outbreak studies were performed between 11 November and 15 December 2020 when the wild-type variant of SARS-CoV-2 was dominant. Clusters of SARS-CoV-2 infections within schools were identified through a prospective school surveillance study. All school contacts of cluster cases, irrespective of symptoms, were invited for PCR testing twice within 48 hrs and 4-7 days later. Combined NTS and saliva samples were collected at each time point along with data on recent exposure and symptoms. Surface and active air samples were collected in the school environment. All samples were PCR-tested and sequenced when possible.

**Results:** Out of 263 sampled school contacts, 24 tested SARS-CoV-2 positive (secondary attack rate 9.1%), of which 62% remained asymptomatic and 42% had a weakly positive test result. Phylogenetic analysis on 12 subjects from 2 schools indicated a cluster of 8 and 2 secondary cases, respectively, but also other distinct strains within outbreaks. Of 51 collected air and 53 surface samples, none were SARS-CoV-2 positive.

**Conclusion:** Our study confirmed within school SARS-CoV-2 transmission and substantial silent circulation, but also multiple introductions in some cases. Absence of air or surface contamination suggests environmental contamination is not widespread during school outbreaks.

## Introduction

The role of schools in the transmission of Severe Acute Respiratory Syndrome Corona Virus 2 (SARS-CoV-2) has been the focus of continuous debate throughout the COVID-19 pandemic. Throughout most of the first year of the pandemic, the Dutch government implemented only a limited set of COVID-19 preventive measures in educational settings to minimise educational disruption. Between November and December 2020, we conducted a series of four detailed outbreak investigations in schools that reported clusters of SARS-CoV-2 infections. At the time, all primary and secondary schools were open and had full occupancy. There was little prior immunity against SARS-CoV-2, vaccines were not yet available and the wild-type variant with the D614G mutation was dominant at the time of the study.

The aim of the outbreak investigations was to provide a more detailed analysis on transmission risk in secondary school settings under the prevailing community incidence and COVID-19 mitigation policy, and to gain insight into the potential role of SARS-CoV-2 transmission via air and surfaces in schools.

## Methods

The outbreak investigations were part of a prospective school surveillance study that evaluates the interactions between indoor air quality, ventilation, environmental SARS-CoV-2 contamination and transmission. At the time the outbreak investigations were performed, physical distancing (>1.5 meters) in schools was implemented for staff-staff and staff-student interactions, but not required in or outside school among children below 18 years. Students and teachers who tested positive for SARS-CoV-2 had to self-isolate at home. Classroom contacts were not quarantined unless considered a close contact, defined as exposure of >15 minutes at <1.5 meter distance to a SARS-CoV-2 infected individual. From December 2020 onwards, mask mandates were in place for students and staff during movement. Seated students and staff did not wear masks. Schools were recommended to increase hand hygiene and the degree of ventilation, plastic shields were installed on teacher desks and all school-based extracurricular activities were cancelled. Testing of asymptomatic close contacts was implemented during the study period (1 December 2020). Other national COVID-19 measures in place at the time, are described in the supplement.

All schools participating in the prospective study kept daily logs of reported SARS-CoV-2 infections among students and staff and whether there was a possible epidemiological link between cases. Schools notified the study team if a cluster was identified, which was defined as three or more cases in the same school within two weeks of whom at least two cases had an epidemiological link. An epidemiological link was defined as cases who shared (class) rooms for at least two course hours in the recent 14 days. If the most recent index case(s) belonging to the cluster had been attending school in the 48 hours prior to symptom onset or PCR test result, an outbreak investigation was initiated.

### Outbreak study

A school visit was scheduled within 48 hours after the notification of a cluster to sample participating school contacts. School contacts were defined as students and staff who had shared a (class)room for at least two course hours in the two days preceding symptom onset in the index case or, if this was unknown, the date of a positive test. A sampling location at the school was set-up where participants could self-collect a combined mid-turbinate NTS sample and a saliva sample under direct supervision of trained study staff after instructions had been provided. For participants who were not present at school, samples were self-collected under supervision of study staff at their home address. Participants also completed a questionnaire including basic demographics, other recent exposure to SARS-CoV-2 infected individuals (other than school index case), prior infection and presence of COVID-19 like symptoms. A second sampling visit was scheduled after 4-7 days, depending on the weekends, for a follow-up NTS and saliva sample from each participant, along with a follow-up questionnaire on recent exposure, symptoms and whether household members tested positive since the previous visit. For a schematic overview of the study design see Supplementary Figure S1. All samples were transported to the laboratory the same day and participants were notified about the results of the PCR test within 48 hours. Positive results were followed by self-isolation as per national policy. Saliva samples were stored at -80? and analysed at a later time.

At the first visit, extensive air and surface sampling took place in school buildings (see Supplementary Methods and Figure S1). Briefly, air samples and surface swab samples were collected at three locations: 1) classrooms attended by students previously in contact with the index cases, 2) the teachers’ lounge and 3) the school cafeteria area. At each location, air sampling consisted of twice a 6-hr filtration-based sample, once a 6-hr cyclone-based sample and once a 1.5-hr impingement-sample (school cafeteria and teachers’ lounge only). Surface swab samples of high- and low-touch surface areas were collected as described previously [1]. A total of five samples were taken in each of the areas above and, when possible, from the classroom where index case teachers were located prior to self-isolation. Field blank samples were collected every other outbreak measurement for air samples, and each outbreak for surface swab samples. Samples were sent to the laboratory at 4 °C and processed within 24-hours.

### Sample analysis

Detailed methods are described in the Supplementary Methods. NTS were collected in tubes containing virus transport medium and total nucleic acid was extracted as described [2]. Oral fluid was collected using the ORACOL S10 saliva collection system (Malvern Medical Developments). Total nucleic acid was extracted using MagNApure 96 (Roche LifeScience) small volume total nucleic acid kit. RT-qPCR was performed as described previously [2], with some modifications on the primers and probe of the RdRP-gene (see Supplementary Methods). From the environmental samples, RNA was isolated using an in-house method as described before [3]. Samples were tested with a SARS-CoV-2 RT-PCR, targeting the E gene of SARS-CoV-2 [2].

Sequencing of NTS RT-PCR positive samples with Ct-values <32 was performed using an amplicon-based approach as described [4]. For RT-PCR positive saliva samples, sequencing was performed using the Nanopore protocol [5,6] with several modifications (see Supplementary Methods for details). The sequences have been submitted to GISAID (www.gisaid.org; accession ID: EPI_ISL_722426-722430, EPI_ISL_722432, EPI_ISL_722290, EPI_ISL_722334.

A secondary case was defined as a school contact participating in the study and testing positive by RT-PCR in at least one of the samples collected during initial or follow-up visits. According to standardised local lab protocols a Ct-value cut-off for sample positivity was set <40 for both targets or at <33 if only one target was positive. Samples were defined as ‘weakly positive’ if the Ct-value for a single target was between 33-40 and negative for the other target.

### Statistical analysis

SARS-CoV-2 incidence rates per schools were calculated by dividing the number of reported infections by the total number of students and staff members. Next, the secondary attack rate (SAR) per cluster was determined by dividing the secondary cases by the total number of participants, both overall and stratified for teachers and students. Case characteristics, school attendance, presence of symptoms and time since exposure were graphically displayed for all secondary cases. For each secondary case, we compared RT-PCR results for NTS versus saliva by means of a paired t-test on the gene with the lowest Ct-value of each specimen type. All successfully sequenced NTS and saliva samples from both test rounds were combined in a phylogenetic reconstruction and are depicted per sample type. If the sequence was available from both test rounds, only the sequence of first round was included in the tree. We also included human sequences from the municipalities of the respective schools as background data, which was retrieved from GISAID. SPSS version 26.0.0.1 (IBM), and R version 4.0.3 (R core team) was used for data management and statistical analysis.

### Ethical statement

The study was not subjected to the Medical Research Involving Human Subjects Act (WMO) and therefore no Ethics review was needed. Informed consent was obtained prior to sampling from participants and, for those <16 years of age, their legal representatives.

## Results

Between 11 November and 15 December 2020, we conducted four outbreak investigations following reports of clusters in four out of the eight schools participating in the prospective study. The overall weekly incidence of SARS-CoV-2 across the four participating schools during the study period varied between 299-820 per 100,000 students and staff members, while the weekly incidence in het Dutch population during the same period varied between 184 and 430 per 100,000 inhabitants (Table 1) [7,8].

**Table 1.**
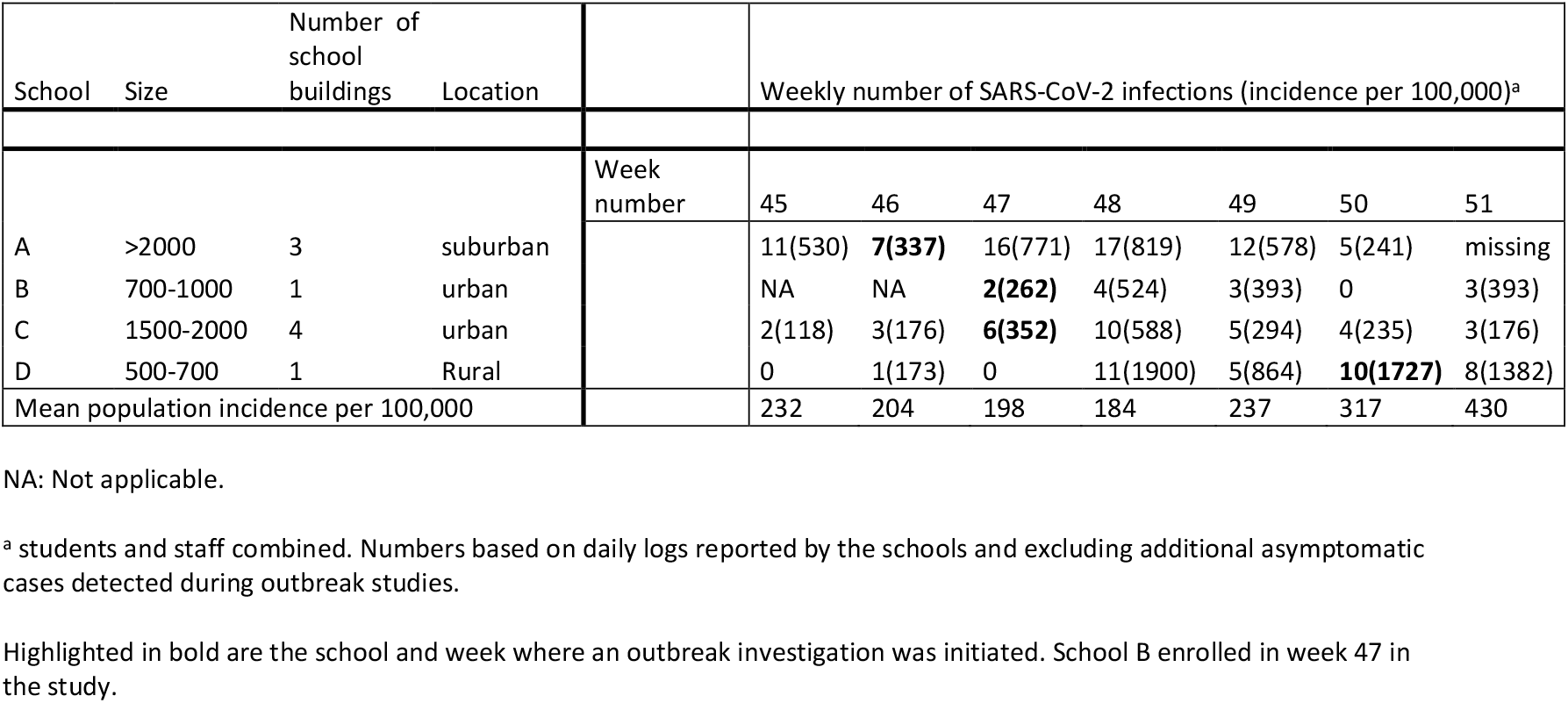
School characteristics and weekly incidence of the four schools participating in the outbreak study (n = 4)

| School | Size | Number of school buildings | Location |  | Weekly number of SARS-CoV-2 infections (incidence per 100,000)<sup>a</sup> |  |  |  |  |  |  |
| --- | --- | --- | --- | --- | --- | --- | --- | --- | --- | --- | --- |
|  |  |  |  | Week number | 45 | 46 | 47 | 48 | 49 | 50 | 51 |
| A | >2000 | 3 | suburban |  | 11(530) | **7(337)** | 16(771) | 17(819) | 12(578) | 5(241) | missing |
| B | 700-1000 | 1 | urban |  | NA | NA | **2(262)** | 4(524) | 3(393) | 0 | 3(393) |
| C | 1500-2000 | 4 | urban |  | 2(118) | 3(176) | **6(352)** | 10(588) | 5(294) | 4(235) | 3(176) |
| D | 500-700 | 1 | Rural |  | 0 | 1(173) | 0 | 11(1900) | 5(864) | **10(1727)** | 8(1382) |
| Mean population incidence per 100,000 |  |  |  |  | 232 | 204 | 198 | 184 | 237 | 317 | 430 |

NA: Not applicable.
<sup>a</sup> students and staff combined. Numbers based on daily logs reported by the schools and excluding additional asymptomatic cases detected during outbreak studies.
Highlighted in bold are the school and week where an outbreak investigation was initiated. School B enrolled in week 47 in the study.

Index cluster size varied between 3 and 12 cases (Supplementary Table S1). In total, 27 SARS-CoV-2 index cases belonged to the clusters, including 6 staff members and 21 students. A total of 1121 school contacts were invited to participate (Figure 1). Cluster C further developed during the outbreak study and additional school contacts were therefore invited during the second sample round. The number of staff exposed to a teacher index case could not be determined reliably as staff-staff contacts occur mostly in the teachers’ lounge. Therefore, all staff members were invited, but informed that they should only participate if they had been in close contact with any of the index cases. In total, 263 school contacts participated, including 93 staff members (Figure 1). The participation rate was 10.6% to 41.8% among staff, and 7.5% to 56.1% among students (Supplementary Table S2). Eighteen subjects participated only in the first test round. In total 508 paired NTS and saliva samples were collected.

**Figure 1.**
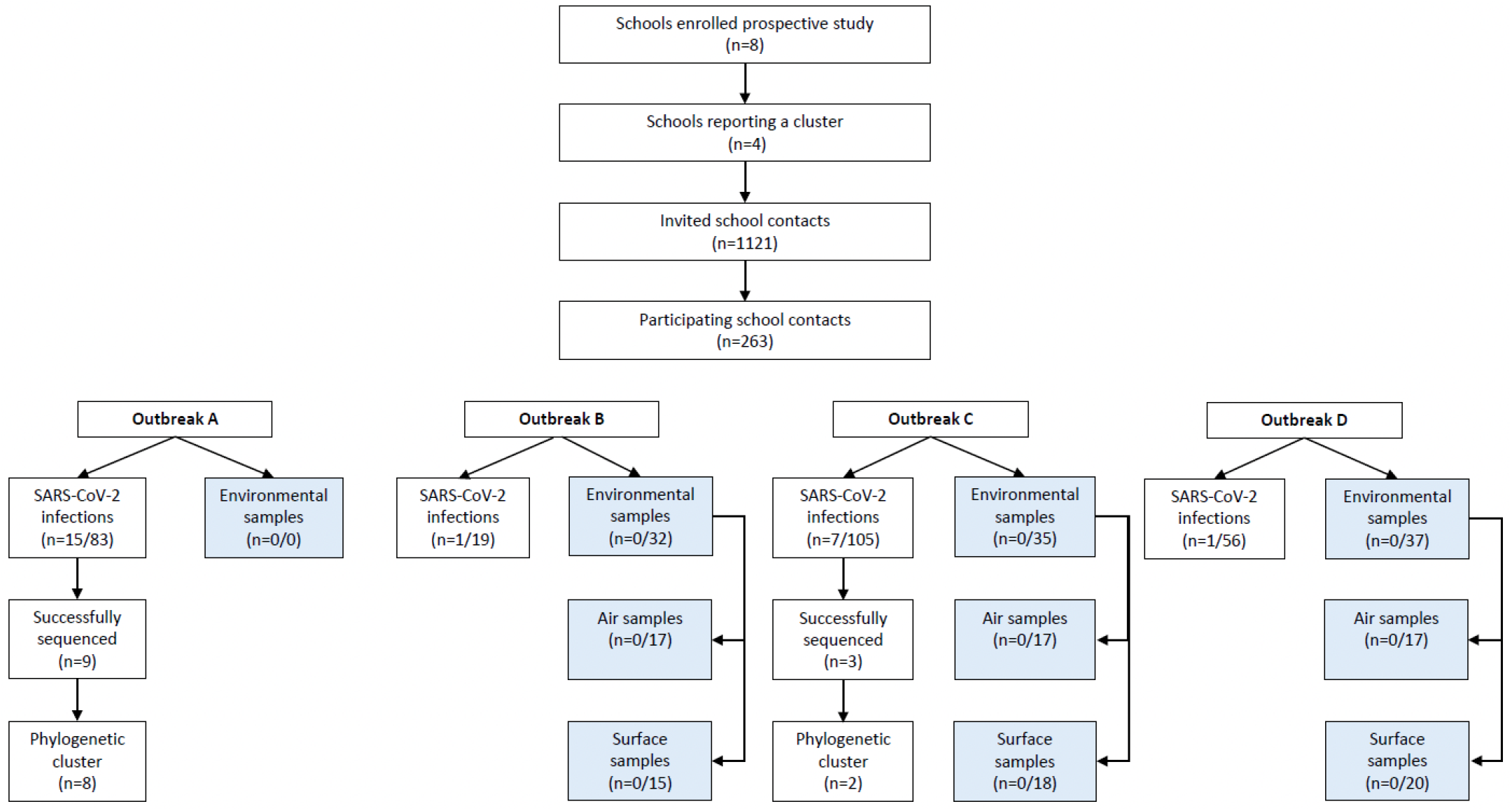
Overview of participating schools and school contacts. Number of SARS-CoV-2 infections among school contacts and positive air and surface samples are depicted per outbreak study. No environmental samples were taken during outbreak A.

### Secondary cases

In total, 24 school contacts (9.1%) tested positive by RT-PCR in at least one sample (Figure 1). Of these, 10 (41.6%) had a weakly positive PCR result. The SAR by cluster varied between 1.8-18.1% and was generally higher among students compared to staff (Supplementary Table S2). Table 2 describes the temporal pattern of exposure, school attendance, SARS-CoV-2 PCR results and symptoms among secondary cases. Out of the 21 secondary cases of whom we obtained symptom data, only eight (38.1%) were symptomatic at any time during follow-up. In four of these participants, the symptoms were present at the time of first sample collection. Notably, three of them attended school while symptomatic. The other four subjects developed symptoms one to three days after the positive PCR result. Out of the 13 asymptomatic individuals, 8 (62%) were weakly positive, while none of the symptomatic individuals were weakly positive. Onwards SARS-CoV-2 infection among household members was reported for 2 out of 21 (10%). One of the participants from school A was already quarantined because of a positive test of a household member, while a student from cluster C reported a family member who tested positive the same day.

**Table 2.**
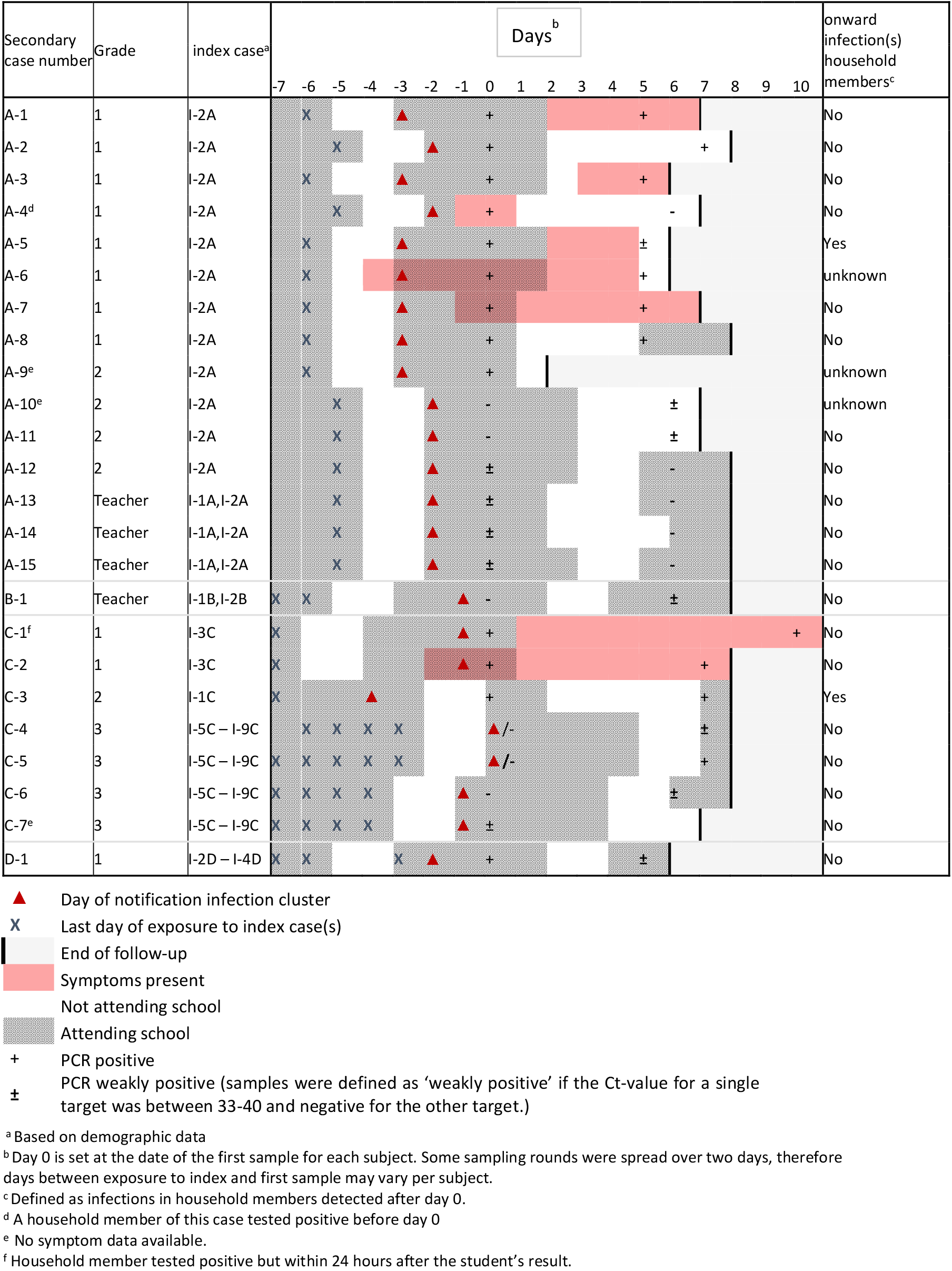
Characteristics of secondary cases belonging to each of the SARS-CoV-2 school clusters (n = 24)

In total 46 paired specimen samples from secondary cases were available (two cases did not participate in the second test round). Discrepancies in test results between the two, self-collected, specimens were observed in 19 out of 46 pairs (Supplementary Table S3). Eight of the 24 secondary cases tested positive only in saliva and five only in NTS. Testing of a second NTS and saliva sample after 3-7 days increased the detection rate by 33%. Lowest Ct-values were detected for samples taken between day five and eight since last exposure and in symptomatic individuals (Figure 2).

**Figure 2.**
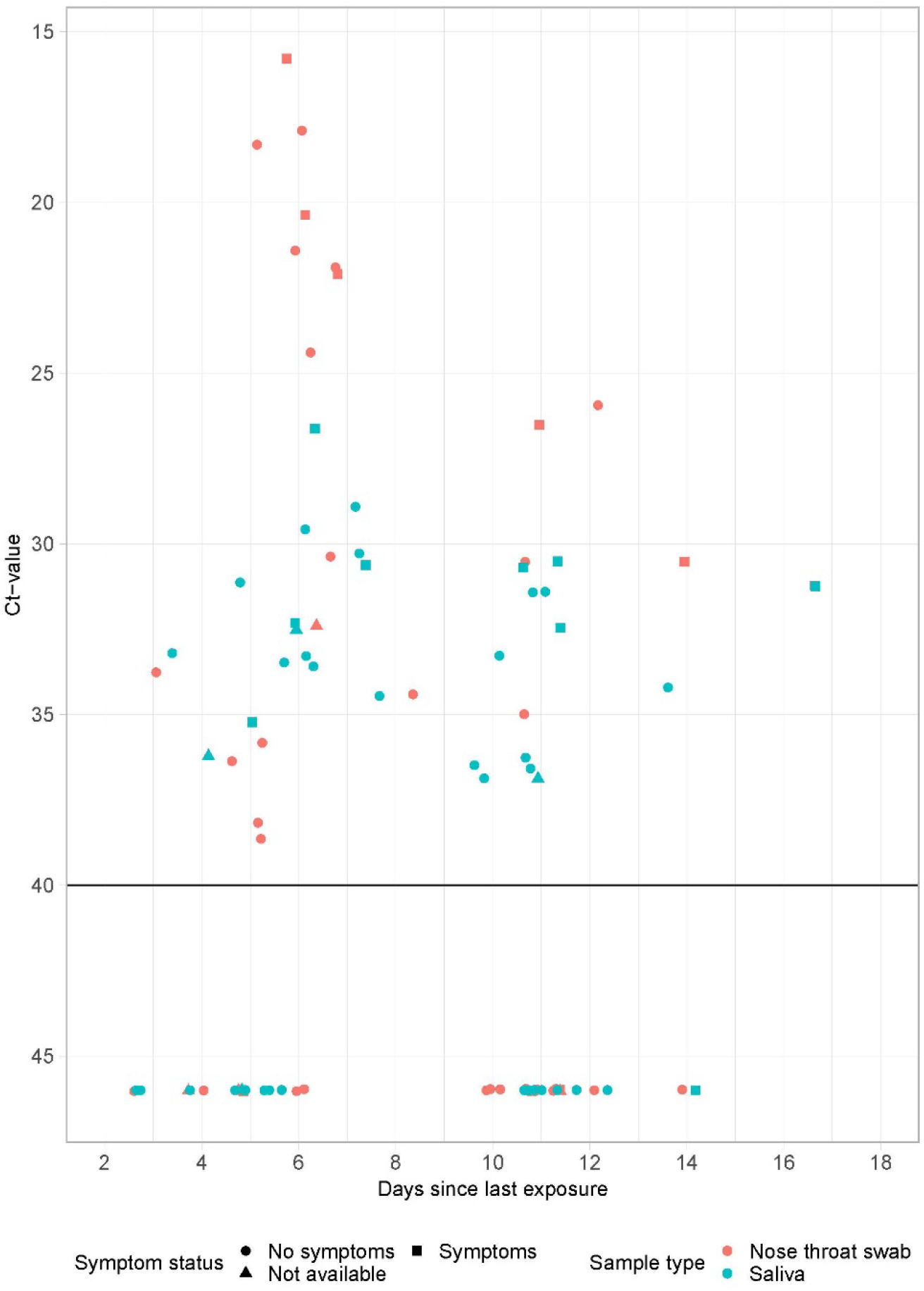
Ct-values by day since last exposure for nose throat swabs and saliva (n = 46) Ct-values by day since last exposure for nose throat swabs (orange) and saliva (blue). All samples from subjects with at least one positive results are included. Negative results are displayed as Ct-value >45.

In school A and C multiple secondary cases were present which allowed to investigate confirmation of a cluster of infection by phylogenetic analysis (Figure 3). A total of 12 individuals were successfully sequenced of which 9 and 3 originated from school A and C respectively.

**Figure 3:**
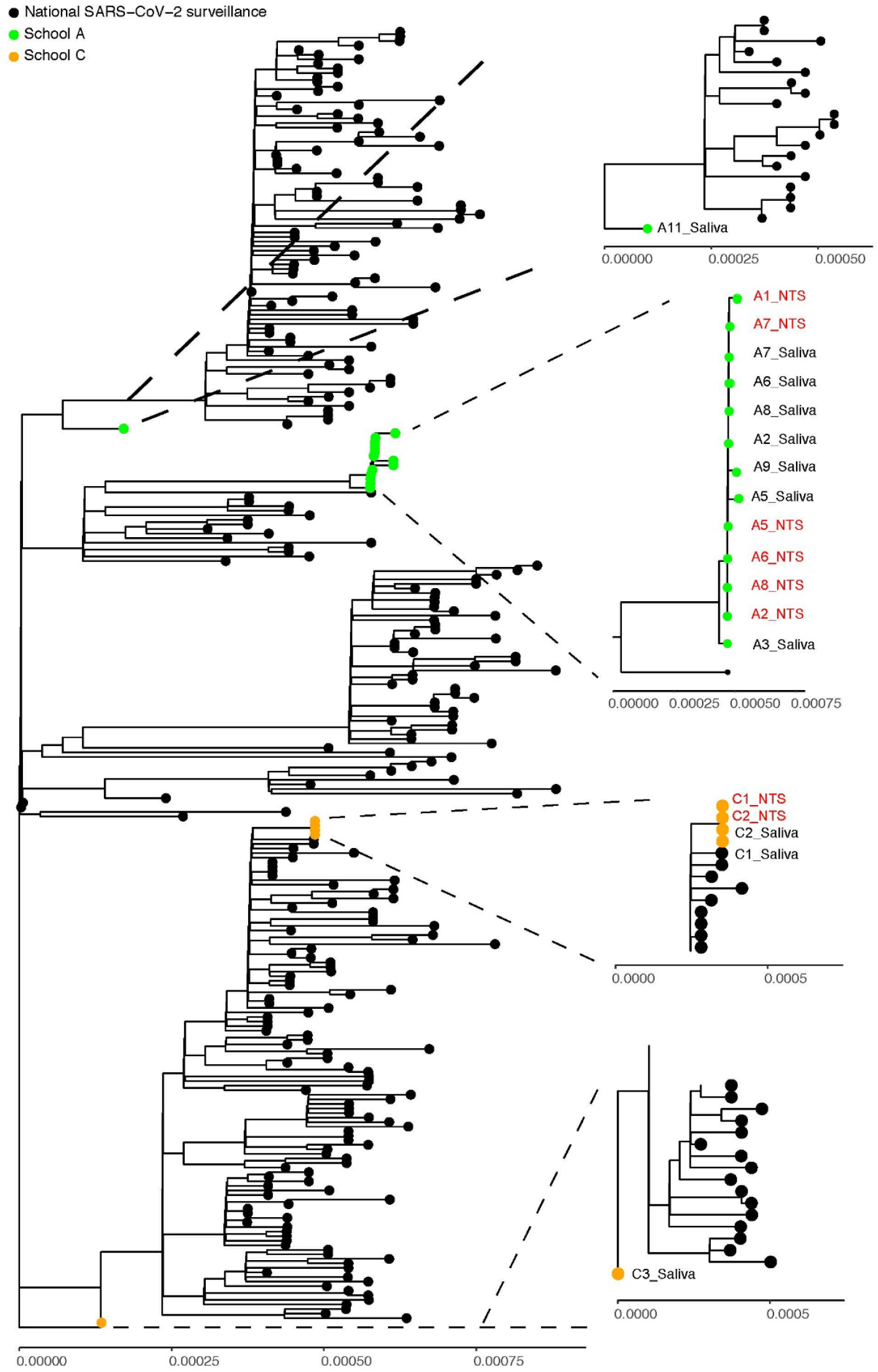
Sequencing data of RT-PCR positive saliva and nose throat swabs within school A and C. NTS: Nose throat swab Both test rounds are included in the figure. If sequencing data was available for both test rounds, only the sequence of the first round is included in the phylogenetic tree. Identification of NTS samples are depicted in red and for saliva samples in black.

In school A, the SARS-CoV-2 RNA sequences originating from eight participants formed a cluster. Demographic data revealed that all participants from this cluster were students and had only been in contact with each other during school hours. They came from two different classes who had been in contact with the same index case teacher. All participants within the cluster did not report any household members or other contacts testing positive before their positive test result. In addition, in one student a divergent strain was identified, indicating a separate instruction.

In school C, four SARS-CoV-2 sequences from two students phylogenetically clustered. They were classmates and only had been in contact with each other and the student index case during classes. One of the students reported a household member who tested positive the same day. For this school, also a phylogenetically distinct strain within the outbreak was identified, again showing a second, independent introduction. These results clearly show genetically linked transmission clusters, but also show other distinct strains within outbreaks, indicative of multiple introductions.

### Environmental sampling of air and surfaces

In total 104 environmental samples were collected from clusters B, C and D at the start of each investigation. All the 51 collected air and 53 surface swab samples tested negative for SARS-CoV-2. All field blank samples tested negative in PCR. For a complete overview of sample locations and results, see Supplementary Results and Table S4.

## Discussion

This detailed outbreak investigation among school contacts of four SARS-CoV-2 clusters using repeated PCR testing of two complementary specimens yielded a positivity rate of 9.1% with the majority of cases (61.9%) being asymptomatic. The sequence results together with detailed contact data indicate the presence of a cluster within school A. Likewise, in school C the sequencing and contact tracing data suggest that two individuals within the same class had infected each other. Combined, these observations indicate that silent transmission of SARS-CoV-2 in secondary schools may occur. Yet, no SARS-CoV-2 contamination was detected in any of the air and/or surface samples collected from the schools during the period of the outbreak, suggesting that environmental contamination was not widespread.

Reported (SAR) values from other school outbreak investigations from the same period that used comparable methodology ranged between 0.0 to 6.5% [9–18]. The majority of the secondary cases in these studies were asymptomatic (47.8%-66.6%), in line with our observations [10,17]. Studies that used symptom based testing alone reported no secondary cases [19,20]. In comparison, in our study symptom-based testing by means of a single NTS as was the policy at the time, would only have identified six cases yielding a SAR of 2.3% compared to 9.1%. In Israel, a large outbreak was reported in a school 10 days after reopening. Testing of the complete school community revealed an attack rate of 13.2% in students and 16.6% in staff members of which 47.8% were asymptomatic [21].

Combined, these results illustrate the importance of the applied testing strategy in estimating outbreak sizes in (school) populations, where silent circulation of SARS-CoV-2 infections can be easily missed. In our study, we increased our detection rate by repeated testing after seven days and combining NTS and saliva samples, which could partially explain our higher detection rate compared to other outbreak studies. It should be noted that 10 individuals were weakly positive and for mitigating transmission (early) detection of such cases may be less important. Nevertheless, the role of asymptomatic and pre-symptomatic infections in propagating the COVID-19 epidemic is now widely acknowledged, in particular among school students because of their more intense contact patterns. In our study, onward transmission to household members was suggested for 10% of the secondary cases for whom this data was available.

Although we found evidence of SARS-CoV-2 transmission within secondary schools, the lack of detectable SARS-CoV-2 RNA in collected air and surface samples suggests that major environmental contamination is uncommon in schools under the prevailing conditions at the time of the study. This is in contrast with findings from previous outbreak investigations conducted at mink farms and nursing homes, where similar sampling technologies were applied [1,22]. In these studies, several air samples collected in COVID-19 infected mink farms, and a high percentage of both air and surface swab samples collected in rooms in nursing homes with SARS-CoV-2 positive patients [1,22]. A previous study in London also found limited evidence of SARS-CoV-2 contamination in school environments [18]. Only in a minority (<2-5%) of surface swab samples taken in both the classrooms of index cases and the washrooms, low amounts of viral RNA could be detected, some of which were collected before deep cleaning took place. In this study only 1/68 (1.5%) of the air samples was positive for SARS-CoV-2 [18].

Several factors could explain our negative results. First, schools implemented various measures to increase (hand) hygiene and prevented social gatherings. Second, schools increased their ventilation regimes by opening doors and windows and installing new mechanical ventilation systems. Although, the effect of these interventions on SARS-CoV-2 transmission is still unclear. Third, in the nursing home and mink farm studies samples were collected in the vicinity of acute phase shedding SARS-CoV-2 patients or minks. This is in contrast with the secondary schools, where the known cases were isolated at home. Moreover, most infected students and teachers were asymptomatic or pauci-symptomatic which is known to be associated with lower infectiousness [23]. Lastly, SARS-CoV-2 spread and transmission is suggested to be a more local phenomenon, suggesting direct droplet contact and/or close range aerosol (up to several meters) as the dominant route of transmission in the school environment [22,24].

The major strength of this study is that we collected a large amount of data in school contacts (e.g. sequencing, symptom onset and recent exposure), irrespective of symptoms, which provided an opportunity to obtain extensive virological and contact tracing information from the subjects. Furthermore, we increased our detection rate by combining specimens and testing twice. Lastly, the combined sampling of the environment and school contacts facilitated identification of transmission mechanisms within secondary schools. However, some limitations need to be addressed: First, we investigated only four outbreaks and observed a high variability in SAR between clusters, reflecting the stochasticity in our data. Second, the low participation rate among contacts may have resulted in under-or overestimation of the SAR due to selection bias. Third, we only invited students to participate if they shared a classroom with the index case. Consequently, we may have missed secondary cases among other school contacts with whom the index case spent time during breaks. Fourth, we cannot conclude that the observed SAR solely reflects school transmission rates, because sequencing of samples was incomplete for secondary cases and not available for index cases. The SAR may therefore have been somewhat inflated by simultaneous unrelated introductions. However, apart from two participants in our study, no other participants reported contact with a known case outside the cluster. Lastly, the outbreak investigation was performed during the pre-alpha period when school aged children were not vaccinated and there was less prior immunity in the population. Therefore, the results should be interpreted in the context of the epidemiological situation at the time.

## Conclusion

In conclusion, our study confirmed within school SARS-CoV-2 transmission, but also multiple introductions and substantial silent circulation at a time with limited COVID-19 prevention measures in secondary school settings and minimal prior immunity. Absence of widespread air or surface contamination suggests transmission may have occurred most likely via direct route or close range aerosol transmission route. Repeated testing is complementary and therefore recommended when complete case detection is desired. These insights can contribute to the discussion on the role of secondary schools in the transmission of SARS-CoV-2 and how to improve future outbreak studies.

## Data Availability

All data produced in the present study are available upon reasonable request to the authors

## Acknowledgement

Authors gratefully acknowledge the participation of the schools in the study. We further thank our colleagues S.Herfst (Erasmus MC, Rotterdam), M.Tersteeg-Zijderveld, J. Spithoven, A.Timan, I. van Schothorst, S. Parga and P. Meijer (IRAS, Utrecht) for their contribution in environmental sample collection, laboratory preparations and sample processing, and laboratory analysis. This study is funded by ZonMw and part of the Scholen en Covid consortium which also involves D.Zhang, E.Ding, M.A. Ortiz-Sanchez and P.M. Bluyssen (TU Delft, Delft). Saliva analyses were financed in part by the Netherlands Ministry of Health, Welfare and Sport. We also gratefully acknowledge the authors, originating and submitting laboratories of the sequences from GISAID’s EpiFlu Database used in the phylogenetic analysis. All contributors of data may be contacted directly via the GISAID website (http://platform.gisaid.org).

## Conflict of interest

None declared

## Authors’ contribution

Lotte Jonker and Kimberly Linde planned and performed human and environmental data collection and performed analyses and drafted the report. Robin Sprado assisted with planning and performing the data collection. Robin Huisman and Richard Molenkamp planned and performed the PCR-testing of the human samples. Bas Oude Munnink and Dirk Eggink planned and performed the sequencing of positive RT-PCR samples. Inge Wouters, Dick Heederik, Pieter Fraaij, Marieke de Hoog and Patricia Bruijning-Verhagen contributed to the overall conceptualization and planning and funding acquisition of the study, and Wietske Dohmen aided in conceptualization of the environmental sampling design All parties contributed to the draft and edits of the report.

## Notes

### Competing Interest Statement

The authors have declared no competing interest.

### Funding Statement

This study was funded by ZonMw. Projectnumber 10430022010024

### Author Declarations

The study was not subjected to the Medical Research Involving Human Subjects Act (WMO). Therefore Medical Research Ethics Committee Utrecht waived ethical approval for this work.

