## Supplementary Materials for "SARS-CoV-2 outbreaks in secondary school settings in the Netherlands during fall 2020; silent circulation"

#### National COVID-19 prevention measures at the time of study

National COVID-19 prevention measures included stay-home orders for persons with respiratory complaints or fever, physical distancing (>1.5 meters) for people ≥ 18 years of age, limiting social gatherings to < 30 people, and a maximum of three visitors per day. Bars and restaurants were closed. Individual sports activities were only allowed outdoors and in teams only for people < 18 years of age. COVID-19 testing was available at municipal health testing facilities for persons with symptoms suspect for COVID-19.

#### Environmental sampling of air and surfaces

At each location, air sampling consisted of twice a filtration-based sample, once a cyclone-based sample, and once a impingement-sample (school cafeteria and teachers' lounge only). The filtration-based technique captures inhalable dust - airborne particles and droplets of an aerodynamic size that enter the respiratory tract through mouth and nose. Air is drawn through an inhalable dust sampling head (Conical Inhalable dust Sampler (CIS), JS Holdings, UK) equipped with 37mm diameter 2.0 µm pore-size Teflon filter (Pall incorporated, Ann Arbor, USA) connected with tubes to a Gilian GilAir 5 pump (Sensidyne, St. Petersburg, USA) calibrated at a flow of 3.5 L/min. The sampling head was attached to a pole at 1.5m height, which is the average breathing height. The sampling train was set up before the beginning of the school day and lasted for 6 hours. In each area, two inhalable dust samplers were placed, one near the teacher's desk and one near the students. After sampling filter holders were detached, they were packed in a MinigripTM bag for transportation at 4 ℃ to the laboratory, and stored at 4 ℃ till further processing next day.

The cyclone-based technique allows for size-selective sampling of dusts and aerosols from the environment. The NIOSH bioaerosol cyclone sampler (NIOSH BC 251, kindly provided to us by William G Lindsley, NIOSH Morgantown, USA) consists of a sampling body which is equipped with a 15ml conical tube (Greiner Bio-One, Alphen aan de Rijn, Netherlands) to capture aerosols sized > 4 µm , a 1.5ml conical tube (Sarstedt BV, Etten-Leur, Netherlands) to capture aerosols with a size ranging between 1-4 µm, and a 37mm diameter filter holder (SKC Incorporated, Eighty Four, USA) equipped with a 37mm diameter 2.0 µm pore-size Teflon filter (Pall incorporated, Ann Arbor, USA) to collect particles of 1 µm and smaller. The filter holder was connected with tubing to a Gilian GilAir 5 pump (Sensidyne, St. Petersburg, USA) calibrated at a flow of 3.5 L/min. The 15ml and 1.5ml tube were pre-filled with 2.5ml and 1.5ml virus transport medium 1 (VTM-1), respectively, before start of the measurement. VTM-1 used for air sampling consisted of HMEM with 25mM Hepes (85 v/v%, Lonza, Verviers Belgium), Hepes 1M (17.5 v/v%), Penicilline/Streptomycine (10000 U/ml, 10000 ug/ml, 2 v/v%, Lonza, Verviers, Belgium), Glycerol 99% grade (10 v/v%), Lactalbumine enymatic hydrolase (0.4 w/v%), Polymyxin B Sulphate (1.67 mg/ml; 1 v/v%), Nystatin (1.69 mg/ml; 0.5 v/v%, Sigma Aldrich, Zwijndrecht, the Netherlands) and gentamycine (0.5 v/v%, Life technologies, Bleiswijk, the Netherlands). The sampling train was set up before the beginning of the school day, and lasted for 6 hours. In each area, one cyclone sampler was placed in conjunction with one of the filter-based samplers. After sampling filter holders were detached, packed in a MinigripTM bag. The 15ml and 1.5ml tubes were detached from the sampling body, subsequently, 1 ml Opti-MEMTM (Gibco, UK) and 1 ml of VTM-1/Opti-MEMTM mixture, respectively, was added to the tubes immediately after sampling. Filter holder and sampling tubes were transported at 4 ℃ to the laboratory, and stored at 4 ℃ till further processing next day.

In the school canteen and teachers' lounge additionally air sampling through impingement was conducted by means of a 5ml BioSampler (SKC Inc, Eighty Four, USA) attached to a pole at 1.5m height. An airflow of 12.5 L/min through the BioSampler was established by connecting the outlet of the sampler to an in-house designed pump unit. The impinger was filled with 4ml of VTM-1 used for air sampling (see above) prior to the beginning of the measurement. The measurement lasted between 1 and 1.5 hour depending on the duration of the break in the corresponding area. Evaporation losses of VTM-1 was replaced by adding every 15 minutes 2 ml of VTM. After sampling, remaining VTM-1 fluid was transferred to a 15ml tube (Greiner BioOne, Etten-Leur, Netherlands) and 2 ml of Opti-MEMTM (Gibco, UK) was added. Tubes containing the collected impinger fluid were transported at 4 ℃ to the laboratory, and stored at 4 ℃ till further processing next day.

Field blank samples one each per type of active air sampling technique were collected every other outbreak measurement.

In each of the above mentioned areas, and if applicable in the classroom where infected teachers provided instructions prior to infection, five swab samples from surfaces were collected. Sampling locations included high touch surfaces like door handles and table tops, and low touch surfaces like top surface of cabinets. To standardise swabbing of surfaces, disposable plastic grids of 10 cm2 were used; when it was not possible to use the grid this was noted. Dry swabs (Medical Wire Dry Swabs, MW370, Corsham, UK) were used, which were placed in 2ml virus transport medium-2 (VTM-2) in 5ml tubes directly after swabbing. VTM-2 used for swabs consisted of DMEM (42.8 v/v %), Hepes (2 v/v%), NaHCO3 (1.2 v/v%), Penicilline/Streptomycine (10000 U/ml; 10000 µg/ml; 10 v/v %) (Lonza, Verviers, Belgium), Amphotericine B (0.25 mg/ml; 4 v/v/%, Eramsus MC Pharmacy, Rotterdam) and Fetal Bovine Serum (40 v/v%, Greiner Bio-One, Etten-Leur, Netherlands). After collection, samples were stored at 4 °C in an electric transport cooling box and transported to the lab. At the lab, samples were placed in 4°C storage until further processing the next day. Field blank swab samples were collected during each outbreak measurement.

#### Laboratory procedures

##### Air and surface samples

All handlings were performed under BSL2+ conditions. RNA was isolated using a previously described in-house method [3] with slight modifications. In short, filters were removed from the filter holders and transferred to 5ml screw-top tubes (Eppendorf, Nijmegen, Netherlands) and 2ml of VTM-1 used for air sampling was added, and subsequently tubes were vortexed for 5 minutes using a vortex adaptor. Tubes containing VTM and OptiMem mixture from NIOSH and impingement samplers were vortexed for 15 seconds, and tubes containing the surface swab samples were vortexed for 1 min prior to further handling. After vortexing 120 µl was transferred and added to a tube containing 180 µl of MagNA Pure 96 External Lysis Buffer (Roche Diagnostics, Almere the Netherlands), followed by 15 seconds of vortexing. VTM/lysis buffer samples were stored frozen at -80 ℃ till transport to Erasmus MC for RT-PCR. A 150 µl VTM/lysisbuffer aliquot was used for RNA extraction as described previously [3]. Presence of SARS-CoV-2 RNA was tested through a SARS-CoV-2 RNA RT-PCR, targeting the E gene of SARS-CoV-2 on Appleid Biosystems 7500 RT PCR, as described previously [2].

##### Nose-throat and saliva samples

NTS were collected in tubes containing 4 mL virus transport medium-3 (Dulbecco's modified eagle's medium (DMEM, Lonza) supplemented with 40% FBS, 20 mM 4-(2-hydroxyethyl)-1-piperazineethanesulfonic acid (HEPES), NaCO3, 10 µg/ml amphotericin B, 1000 U/mL penicillin, 1000 µg/mL streptomycin). Samples were shipped and processed the same day at the laboratory of the Erasmus Medical Center. Total nucleic acid was extracted from 500 µl NTS fluid using MagNApure 96 (Roche LifeScience) total nucleic acid kit with external lysis protocol and eluted in 100 ul. RT-PCR detection of SARS-CoV-2 was performed as described [2].
Oral fluid was collected using the ORACOL S10 saliva collection system (Malvern Medical Developments). Samples were immediately cooled and shipped to the laboratory of the National Institute of Public Health (RIVM) the same day, where they were stored at -80C until further processing. Total nucleic acid was extracted from 200 µl oral fluid using MagNApure 96 (Roche LifeScience) small volume total nucleic acid kit and elution in 50 µl. RT-qPCR was performed on 5 µl extract using TaqMan® Fast Virus 1-Step Master Mix (Thermo Fisher) on Roche LC480II thermal cycler with SARS-like beta coronavirus (Sarbeco) specific E-gene primers and probe as described previously with following modifications [2]. The primers and probe of the RdRP-gene SARS-CoV-2 RT-PCR have been modified from the original Corman primers and probe conferring the primers SARS-CoV-2 specific and the analytical sensitivity comparable to that of the E-gen RT-qPCR. Modified primers and probe: RdRp_SARS-F2 GTGAAATGGTCATGTGTGGCGG; RdRp_SARS-R2 CAAATGTTAAAAACACTATTAGCATAAGCA; RdRp_SARS-P2.2 CCAGGTGGAACCTCATCAGGAGATGC.

#### Sequencing of NTS and saliva samples

Sequencing of NTS RT-PCR positive samples with Ct-values <32 was performed using an in house designed amplicon-based approach as described before [4].

For RT-PCR positive saliva samples sequencing was performed using the Nanopore protocol "PCR tiling of COVID-19 virus (Version: PTC_9096_v109_revE_06FEB2020)" which is based on the ARTIC v3 amplicon sequencing protocol [5,6]. Several modifications were made to the protocol as primer concentrations were increased from Both libraries were generated using native barcode kits from Nanopore SQK-LSK109 (EXP-NBD104, EXP-NBD114 and EXP-NBD196) and sequencing was performed on a R9.4.1 flow cell multiplexing 48 or 96 samples per sequence run.

**Figure S1.** Overview of study procedures during an outbreak investigation at a school. Environmental samples are depicted in blue and human samples are depicted in white

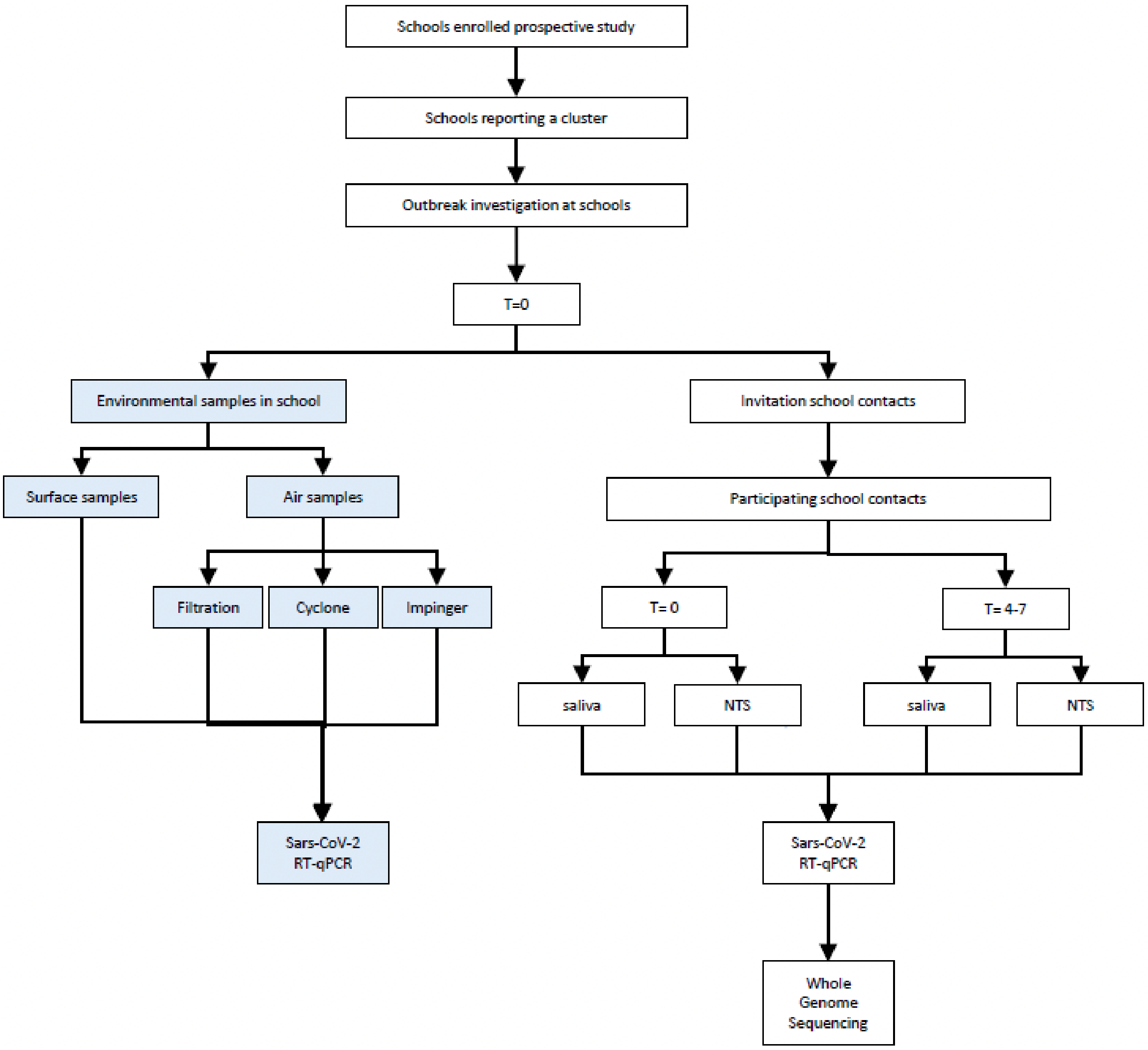

### Supplementary results

**Table S1. Characteristics of index cases**

| Index case | Staff/student | Days since 1st index[a] | Days at school[b] | Any symptoms |
|---|---|---|---|---|
| I-1A | Staff | 0 | 1 | Y |
| I-2A | Staff | 0 | 1 | Y |
| I-3A | Student | 2 | 1 | Y |
| I-1B | Staff | 6 | 2 | Y |
| I-2B | Staff | 7 | 1 | Y |
| I-3B | Student | 0 | 1 | Y |
| I-1C | Staff | 1 | 2 | Y |
| I-2C | Student | 0 | 1 | Y |
| I-3C | Student | 0 | 1 | Y |
| I-4C | Student | 0 | 1 | Y |
| I-5C | Student | 11 | 2 | Unknown |
| I-6C | Student | 10 | 2 | Y |
| I-7C | Student | Unkown | Unknown | Y |
| I-8C | Student | 9 | 2 | Y |
| I-9C | Student | 13 | 2 | Unknown |
| I-1D | Student | 2 | 3 | Unknown |
| I-2D | Student | 0 | 2 | Y |
| I-3D | Student | 3 | 0 | N |
| I-4D | Student | 4 | 8 | Y |
| I-5D | Staff | 6 | 0 | Y |
| I-6D | Student | 9 | 1 | Y |
| I-7D | Student | 8 | 3 | Y |
| I-8D | Student | 7 | 1 | Unknown |
| I-9D | Student | 9 | 1 | Y |
| I-10D | Student | 9 | 1 | Y |
| I-11D | Student | 9 | Unknown | Unknown |
| I-12D | Student | 10 | 3 | Y |

[a] Number of days between positive test in current case and first index case in the cluster
[b] Days at school; number of days present at school during the presumed infectious period (i.e. from two days before date of symptom onset or, if unknown/asymptomatic, the test date)

**Table S2. Details of outbreak investigations**

| Cluster | | Invited | Participated | Median age participants (IQR) | Percentage male | SARS-CoV-2 positive | positivity rate |
|---|---|---|---|---|---|---|---|
| A | staff | 184 | 28 (15.2%) | 33.7 (29.8-55.1) | 46.4% | 3 | 10.7% |
| | students | 98 | 55 (56.1%) | 13.0 (12.4-13.6) | 20.0% | 12 | 21.8% |
| B | staff | 104 | 11 (10.6%) | 40.1 (31.5-48.3) | 63.6% | 1 | 9.1% |
| | students | 106 | 8 (7.5%) | 12.7 (12.3-13.6) | 50.0% | 0 | 0.0% |
| C | staff | 182 | 26 (14.3%) | 41.9 (36.2-57.4) | 50.0% | 0 | 0.0% |
| | students | 241 | 79 (32.8%) | 15.3 (14.8-15.7) | 44.3% | 7 | 8.9% |
| D | staff | 67 | 28 (41.8%) | 49.3 (40.5-58.8) | 35.7% | 0 | 0.0% |
| | students | 139 | 28 (20.1%) | 12.9 (12.7-14.7) | 57.1% | 1 | 3.6% |

**Table S3. Ct-values of different specimens and sample time points among secondary cases (n=24)**

| First sampling round | | | | | | Second sampling round | | | | |
|---|---|---|---|---|---|---|---|---|---|---|
| Secondary case | Days since last exposure | Nose throat swab Ct-value | | Saliva Ct-value | | Days since last exposure | Nose throat swab Ct-value | | Saliva Ct-value | |
| | | RdRP-gen | E-gen | RdRP-gen | E-gen | | RdRP-gen | E-gen | RdRP-gen | E-gen |
| A-1 | 6 | **27.30** | **24.40** | **33.59** | **34.49** | 11 | **26.50** | **26.50** | **32.46** | **35.85** |
| A-2 | 5 | **20.80** | **18.30** | **31.17** | **31.13** | 12 | **28.90** | **25.90** | Neg | Neg |
| A-3 | 6 | Neg | Neg | **33.29** | **35.28** | 11 | Neg | Neg | **30.51** | **31.52** |
| A-4 | 5 | Neg | Neg | **35.22** | **35.22** | 11 | Neg | Neg | Neg | Neg |
| A-5[a] | 6 | **23.90** | **21.40** | **33.51** | **33.48** | 11 | Neg | Neg | Neg | **36.26** |
| A-6 | 6 | **22.30** | **20.40** | **27.02** | **26.63** | 11 | Neg | **30.50** | **31.89** | **31.42** |
| A-7 | 6 | **19.60** | **15.80** | **32.95** | **32.31** | 11 | Neg | Neg | **30.69** | **31.70** |
| A-8 | 6 | **20.60** | **17.90** | **29.95** | **29.58** | 11 | Neg | **35.00** | **31.40** | **32.76** |
| A-9[a] | 6 | Neg | **32.40** | **32.52** | **33.02** | *NA* | - | - | - | - |
| A-10[a] | 5 | Neg | Neg | Neg | Neg | 11 | Neg | Neg | Neg | **36.88** |
| A-11 | 5 | Neg | Neg | Neg | Neg | 11 | Neg | Neg | Neg | **36.58** |
| A-12 | 5 | Neg | **35.80** | Neg | Neg | 11 | Neg | Neg | Neg | Neg |
| A-13 | 5 | Neg | **38.20** | Neg | Neg | 11 | Neg | Neg | Neg | Neg |
| A-14 | 5 | Neg | **38.60** | Neg | Neg | 11 | Neg | Neg | Neg | Neg |
| A-15 | 5 | Neg | **36.40** | Neg | Neg | 11 | Neg | Neg | Neg | Neg |
| B-1 | 6 | Neg | Neg | Neg | Neg | 12 | Neg | **36.60** | Neg | Neg |
| C-1 | 7 | **24.30** | **21.90** | **30.76** | **30.28** | 17 | **33.70** | **31.20** | **31.24** | **31.51** |
| C-2 | 7 | **24.50** | **22.10** | **30.62** | **31.93** | 14 | **31.60** | **30.50** | Neg | Neg |
| C-3 | 7 | **32.10** | **30.40** | **28.91** | **29.18** | 14 | Neg | Neg | **34.21** | **34.84** |
| C-4 | 3 | Neg | Neg | Neg | Neg | 10 | Neg | Neg | Neg | **36.48** |
| C-5 | 3 | Neg | Neg | Neg | Neg | 10 | Neg | Neg | **33.28** | **35.31** |
| C-6 | 4 | Neg | Neg | Neg | Neg | 10 | Neg | Neg | Neg | **36.87** |
| C-7[a] | 4 | Neg | Neg | Neg | **36.22** | *NA* | - | - | - | - |
| D-1 | 3-7 | Neg | **33.80** | **33.20** | **33.52** | 8-12 | Neg | **34.40** | Neg | **34.46** |

[a] Case had symptoms between day 3-5, but not on test days
[b] No symptom data available

Highlighted in red are samples from individuals who reported symptoms at the day of the test

**Table S4. Descriptives and results of detection of SARS-CoV-2 RNA in air and surface samples in the school environment during outbreak measurements.**

| Outbreak | Location | Air measurements | | | | | Surface measurements |
|---|---|---|---|---|---|---|---|
| | | CIS | NIOSH | | | Impinger | Surface swab |
| | | # pos /n | > 4 µm # pos /n | 1-4 µm # pos /n | ≤ 1 µm # pos /n | # pos /n | # pos /n |
| A[a] | Occupied classroom | | | | | | |
| | Teachers' lounge | | | | | | |
| | Canteen | | | | | | |
| | Index case classroom | | | | | | |
| B | Occupied classroom | 0/2 | 0/1 | 0/1 | 0/1 | n.a. | 0/5 |
| | Teachers' lounge | 0/2 | 0/1 | 0/1 | 0/1 | 0/1 | 0/5 |
| | Canteen | 0/2 | 0/1 | 0/1 | 0/1 | 0/1 | 0/5 |
| | Index case classroom | n.a. | n.a. | n.a. | n.a. | n.a. | n.a. |
| C | Occupied classroom | 0/2 | 0/1 | 0/1 | 0/1 | n.a. | 0/4 |
| | Teachers' lounge | 0/2 | 0/1 | 0/1 | 0/1 | 0/1 | 0/4 |
| | Canteen | 0/2 | 0/1 | 0/1 | 0/1 | 0/1 | 0/5 |
| | Index case classroom | n.a. | n.a. | n.a. | n.a. | n.a. | 0/5 |
| D | Occupied classroom | 0/2 | 0/1 | 0/1 | 0/1 | n.a. | 0/5 |
| | Teachers' lounge | 0/2 | 0/1 | 0/1 | 0/1 | 0/1 | 0/5 |
| | Canteen | 0/2 | 0/1 | 0/1 | 0/1 | 0/1 | 0/5 |
| | Index case classroom | n.a. | n.a. | n.a. | n.a. | n.a. | 0/5 |
| Total | | 0/18 | 0/9 | 0/9 | 0/9 | 0/6 | 0/53 |

n: total number of samples.
Pos: Positive result; Cut-off of Ct<40.
n.a.: no air samples collected in the infected classroom.

[a] During the first outbreak investigation, the environmental samples were not collected.

Occupied classroom = classroom where students who have previously been in contact with the index case(s) were instructed.
Index classroom = classroom where infected teachers instructed before going into quarantine.
